# COVID-19 Vaccine Booster Dose Willingness Among Patients with Inflammatory Bowel Disease on Infliximab and Vedolizumab: A Cross-Sectional Study

**DOI:** 10.1101/2022.07.18.22277757

**Authors:** Mohammad Shehab, Fatema Alrashed, Ahmad Alfadhli

## Abstract

**Background:** Vaccination has been effective in preventing COVID-19 infections and related mortality. However, waning immunity after the two-dose vaccination prompted health authorities to recommend a third dose of COVID-19 vaccine to boost immunity. The aim of our study was to assess willingness to receive a third (booster) dose among patients with IBD.

**Methods:** A cross-sectional study was performed at a tertiary care inflammatory bowel disease center. Patients were recruited at the infusion room from January 1st, 2022, until March 31st, 2022. The primary outcome was the prevalence of BNT162b2 third (booster) dose in infliximab- or vedolizumab-treated patients with IBD. The secondary outcome evaluated whether the prevalence of BNT162b2 third (booster) dose differed based on type of COVID-19 vaccine, gender, age, type of biologic therapy and citizenship.

**Results:** In total, 499 patients with IBD were included in this study. The median age was 34.5 years, and 60% had ulcerative colitis (UC). Among the study participants, 302 (60.5%) patients were vaccinated with BNT162b2, and 197 (39.5%) were vaccinated with ChAdOx1 nCoV-19. Of the total number of participants, 400 (80.2%) were receiving infliximab, and 99 (19.8%) were receiving vedolizumab. Overall, 290 (58.1%) of the included patients were willing to receive the third (booster) dose. Patients vaccinated with BNT162b2 were more likely to receive booster dose compared to patients vaccinated with ChAdOx1 nCoV-19 [201 (66.5%) vs 101 (33.5%), p = 0.014]. Infliximab-treated patients were more likely to receive booster dose compared to patients receiving vedolizumab [310 (77.5%) vs 62 (62.6%), p = 0.002]. There was no statistical difference in willingness to receive booster dose in terms of age, nationality, or gender.

**Conclusion:** The percentage of patients with IBD willing or have received a third (booster) dose of BNT162b2 vaccine was lower compared to general population. In addition, patients who received two doses of BNT162b2 vaccines were more likely to receive a third (booster) dose compared to patients who received ChAdOx1 nCoV-19. Patients treated with infliximab were more likely to receive a third (booster) dose of COVID-19 vaccine.

## 1. Introduction

The global spread of COVID-19 has led to multiple vaccines available and approved for use. Typically, the severe acute respiratory syndrome Coronavirus 2 (SARS-CoV-2) vaccines compromise of two dose series to generate full immunity, however, as new variants emerge, uncertainty about duration of immunity after the first two doses and possible risk of breakthrough infection led to approval of a third (booster) dose of a COVID-19 vaccine.^1^

Patients with inflammatory bowel diseases (IBDs), namely Crohn’s disease (CD) and ulcerative colitis (UC), often require immune-modifying treatment, which might increase the risk of opportunistic infection,^2^ therefore, routine check of their vaccination history is performed at diagnosis or before immune suppressive treatment is started. Despite the increased risk of opportunistic infections with some immune suppressive therapy, growing body of evidence have demonstrated that IBD patients were not found to be at an increased risk of developing Coronavirus disease 2019 (COVID-19) or of experiencing a more severe disease.^3,4^ In addition, studies have showed that antibody response post-COVID-19 vaccination is robust in patients with IBD treated with immunosuppressive therapy,^5,6^ therefore, vaccination is strongly encouraged in patients with IBD as the benefits of immunization outweigh the risks or the possibility of suboptimal response.^7,8^

In February 2020, Kuwait reported the first confirmed case of COVID-19, and in April 2020 the first death due to COVID-19 was announced.^9^ Vaccination was initiated in December 2020. BNT162b2 and ChAdOx1 nCoV-19 vaccines were the only available vaccines initially, at later stages of the pandemic mRNA-1273 was also approved. However, BNT162b2 vaccine is the only approved booster vaccine in Kuwait. As of 22 May 2022, a total of 8,022,755 vaccine doses have been administered.^9^ A previous study published in December 2021 explored the prevalence of COVID-19 vaccine in patients with IBD on biologic therapies in Kuwait.^10^ The study found that compared to the general population, the overall prevalence of COVID-19 vaccination in patients with IBD was lower.

Evidence of the safety and efficacy of a third (booster) dose of SARS-CoV-2 vaccine is growing.^8,11^ Reassuringly, the majority of currently available survey studies have reported a high willingness among patients with IBD to get vaccinated against SARS-CoV-2.^12^ However, willingness to receive third (booster) dose in patients with IBD is still unknown. The aim of our study was to assess willingness to receive a third (booster) dose of BNT162b2 among patients with IBD in Kuwait and to evaluate possible factors associated with unwillingness to receive a third (booster) dose of COVID-19 vaccine.

## 2. Material and Methods

A cross-sectional study was performed at a tertiary care inflammatory bowel disease center, Mubarak Al-Kabeer University Hospital, in Kuwait. Patients were recruited at the gastroenterology infusion room from January 1^st^, 2022 until March 31^st^, 2022. Third (booster) dose SARS-CoV2 vaccination counseling was performed by health care providers during clinic and infusion room visits before recruitment. Eligibility criteria included patients: (1) with confirmed diagnosis of inflammatory bowel disease before the start of the study, (2) who received two doses of SARS-CoV-2 vaccine before the start of the study, (3) were on infliximab or vedolizumab at least six weeks prior to recruitment, (4) are 18 years of age or older. We excluded patients with: (1) confirmed or suspected SARS-CoV-2 infection after the second dose of vaccination, (2) are on concurrent corticosteroids use within two weeks of the start of the study, (3) Developed severe adverse effects from previous SARS-CoV-2 vaccination dose. Reliability was performed by evaluating vaccination status of participants in two different visits to the infusion room, with eight weeks separating each visit. Electronic medical records of included patients were used to obtain disease characteristics, demographics, and vaccination details. The study was performed and reported in accordance with the Strengthening the Reporting of Observational Studies in Epidemiology (STROBE) guidelines.^13^

The primary outcome was the percentage of patients willing or have received a third (booster) dose of BNT162b2 vaccination among patients with IBD on infliximab or vedolizumab. The secondary outcome evaluated whether the prevalence of BNT162b2 third (booster) dose differed based on type of COVID-19 vaccine, gender, age, type of biologic therapy and citizenship.

The international classification of diseases (ICD-10 version: 2019) was used to make diagnosis of IBD. Patients were considered to have IBD when they had ICD-10 K50, K50.1, K50.8, K50.9 indicating Crohn’s disease (CD) or ICD-10 K51, K51.0, K51.2, K51.3, K51.5, K51.8, K51.9 indicating ulcerative colitis (UC).^14^

Descriptive analyses were conducted to calculate frequencies and proportions of categorical variables. Chi-squared test (X^2^) to evaluate the difference in willingness to receive third (booster) dose differed across categories of demographic variables. The statistical significance level was set at α = 0.05. All Analyses were conducted using R (R core team, 2017).

## 3. Results

Between January 1st, 2022, until February 28th, 2022, 575 patients were interviewed, of which 76 patients were excluded because of confirmed SARS CoV-2 infection after second dose of vaccine. Therefore, 499 patients with inflammatory bowel disease (IBD) were included in this study. The median age was 34.5 years, and more than half of the included patients were female (54.9%). 40% of included patients had Crohn’s disease (CD) and 60% had ulcerative colitis (UC). The median BMI was 25.1 kg/m^2^, and 109 (22.0%) of the participants were smokers. Asthma (11.2%), diabetes (8.0%), and hypertension (6.6%) were the most reported co-morbidities among the study participants. The median duration of infliximab and vedolizumab therapy at the time of the study was 12 and 11 months respectively. (Table 1).

**Table 1.**
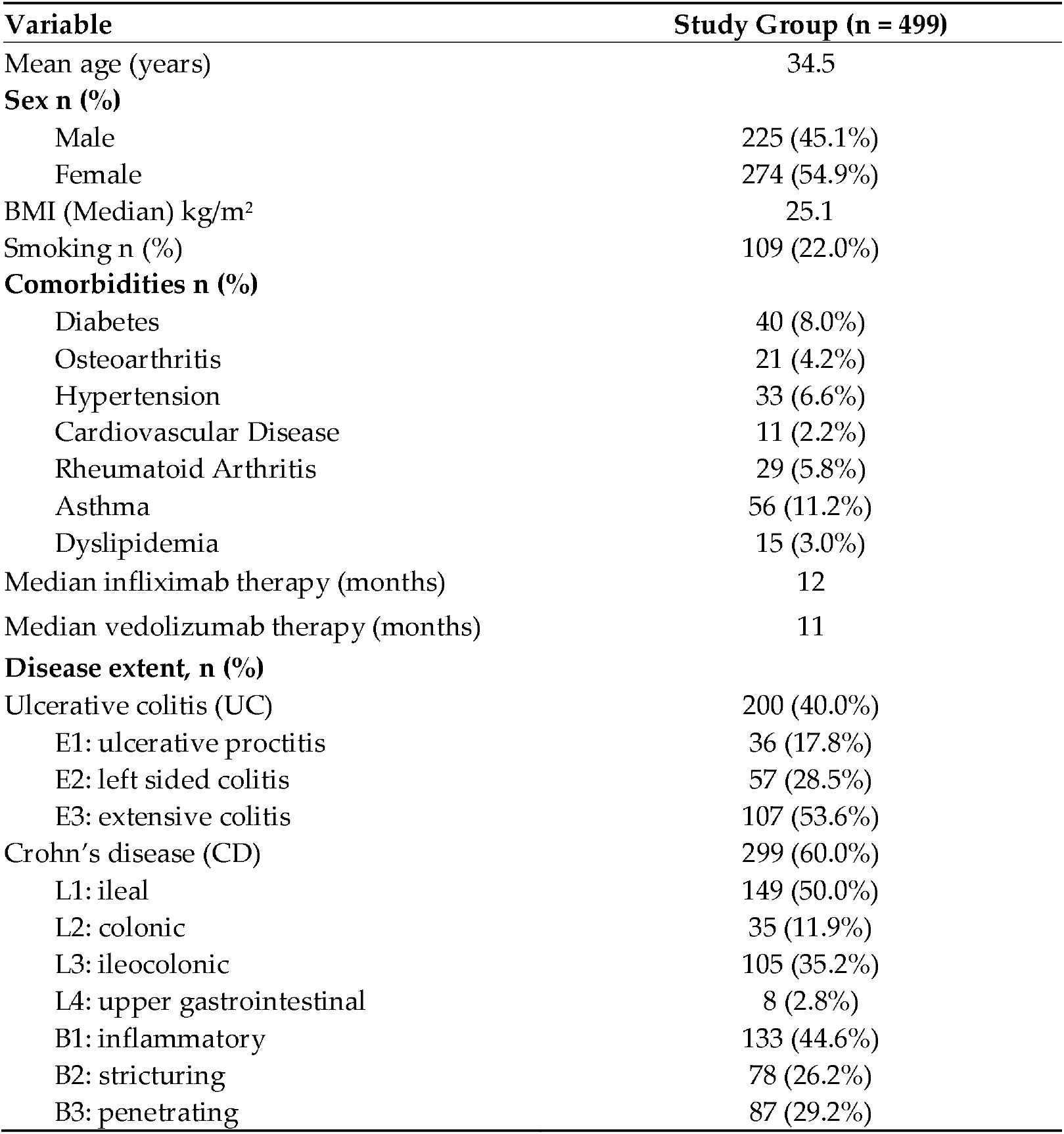
Demographics of Pa<u>tients with IBD.</u>

| Variable | Study Group (n = 499) |
| --- | --- |
| Mean age (years) | 34.5 |
| **Sex n (%)** |  |
| Male | 225 (45.1%) |
| Female | 274 (54.9%) |
| BMI (Median) kg/m<sup>2</sup> | 25.1 |
| Smoking n (%) | 109 (22.0%) |
| **Comorbidities n (%)** |  |
| Diabetes | 40 (8.0%) |
| Osteoarthritis | 21 (4.2%) |
| Hypertension | 33 (6.6%) |
| Cardiovascular Disease | 11 (2.2%) |
| Rheumatoid Arthritis | 29 (5.8%) |
| Asthma | 56 (11.2%) |
| Dyslipidemia | 15 (3.0%) |
| Median infliximab therapy (months) | 12 |
| Median vedolizumab therapy (months) | 11 |
| **Disease extent, n (%)** |  |
| Ulcerative colitis (UC) | 200 (40.0%) |
| E1: ulcerative proctitis | 36 (17.8%) |
| E2: left sided colitis | 57 (28.5%) |
| E3: extensive colitis | 107 (53.6%) |
| Crohn's disease (CD) | 299 (60.0%) |
| L1: ileal | 149 (50.0%) |
| L2: colonic | 35 (11.9%) |
| L3: ileocolonic | 105 (35.2%) |
| L4: upper gastrointestinal | 8 (2.8%) |
| B1: inflammatory | 133 (44.6%) |
| B2: stricturing | 78 (26.2%) |
| B3: penetrating | 87 (29.2%) |

Among the study participants, 302 (60.5%) patients were vaccinated with BNT162b2, and 197 (39.5%) were vaccinated with ChAdOx1 nCoV-19. Of the total number of participants, 400 (80.2%) were receiving infliximab, and 99 (19.8%) were receiving vedolizumab. Among infliximab-treated patients, 275 (68.7%) patients were receiving concomitant immunosuppressive therapy.Regarding age, 100 (20.0%) participants were above the age of 40, and 399 patients (80.0%) were age 40 or above. In terms of nationality, 346 (69.3%) of participants were Kuwaitis and 153 (30.7%) were expatriates.

Overall, 290 (58.1%) of the included patients were willing to receive the third (booster) dose. Patients with IBD vaccinated with BNT162b2 were more likely to receive booster dose compared to patients vaccinated with ChAdOx1 nCoV-19 [201 (66.5%) vs 101 (33.5%), p = 0.014](Figure1). In terms of biologic therapy, patients receiving infliximab were more likely to receive booster dose compared to patients receiving vedolizumab [310 (77.5%) vs 62 (62.6%), p = 0.002](Figure 2). There was no statistical difference in willingness to receive booster dose in terms of age, nationality or gender. (Table 2)

**Figure 1.**
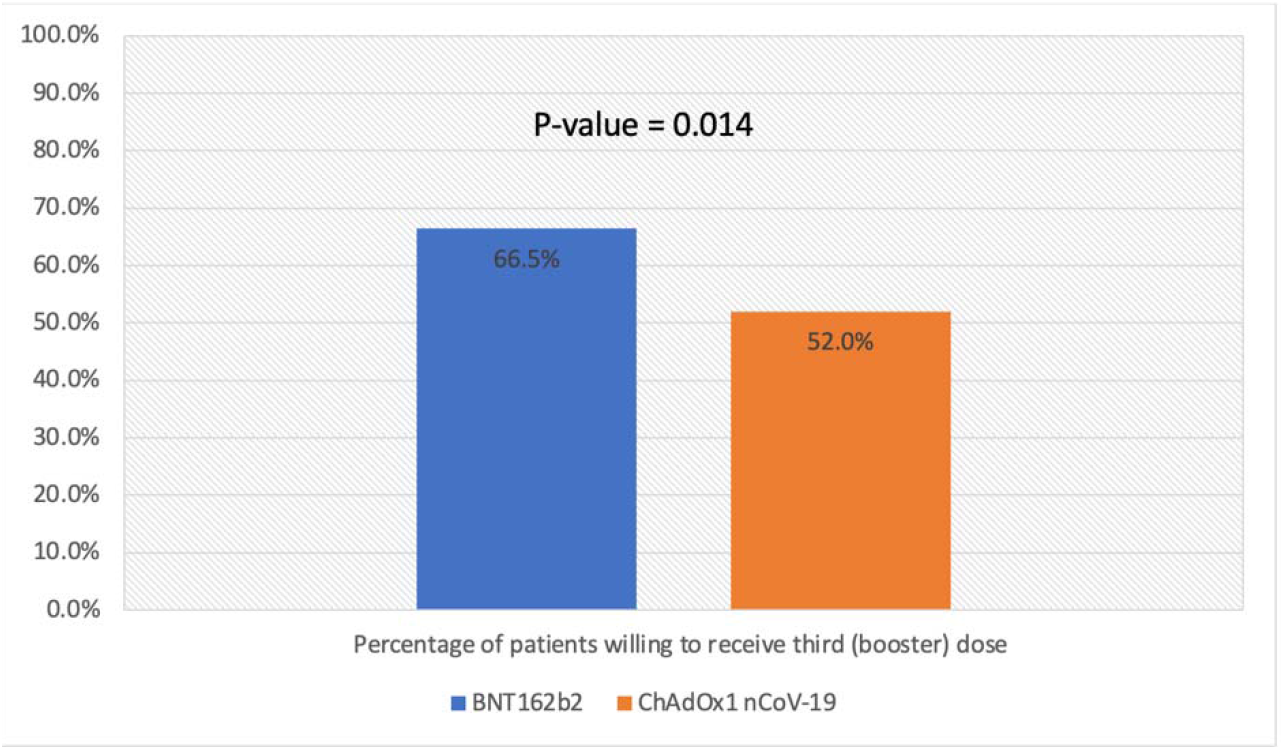
Percentage of patients willing to receive third (booster) dose stratified by type of previous vaccine.

**Table 2.**
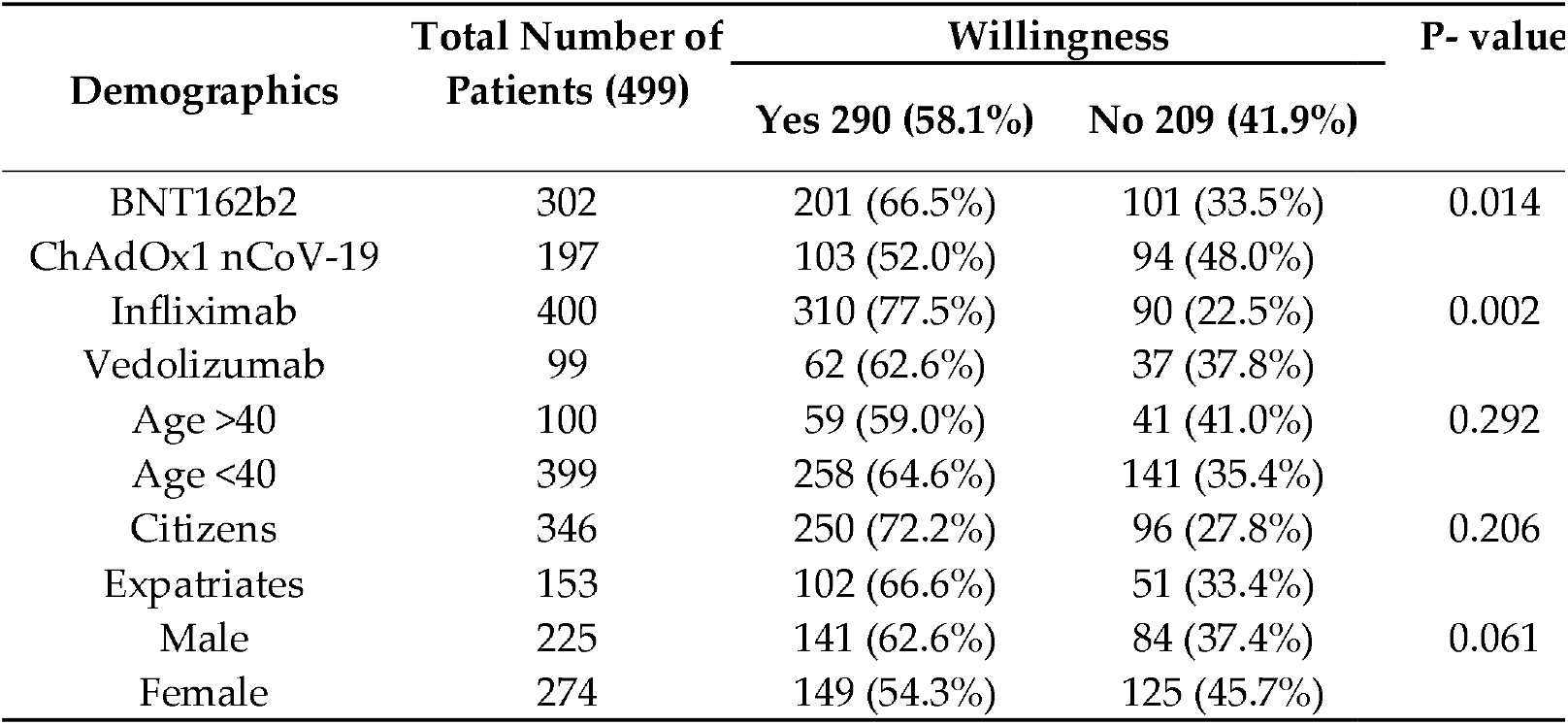
Demographics of patients with IBD according to willingness.

| Demographics | Total Number of Patients (499) | Willingness |  | P- value |
| --- | --- | --- | --- | --- |
|  |  | Yes 290 (58.1%) | No 209 (41.9%) |  |
| BNT162b2 | 302 | 201 (66.5%) | 101 (33.5%) | 0.014 |
| ChAdOx1 nCoV-19 | 197 | 103 (52.0%) | 94 (48.0%) |  |
| Infliximab | 400 | 310 (77.5%) | 90 (22.5%) | 0.002 |
| Vedolizumab | 99 | 62 (62.6%) | 37 (37.8%) |  |
| Age >40 | 100 | 59 (59.0%) | 41 (41.0%) | 0.292 |
| Age <40 | 399 | 258 (64.6%) | 141 (35.4%) |  |
| Citizens | 346 | 250 (72.2%) | 96 (27.8%) | 0.206 |
| Expatriates | 153 | 102 (66.6%) | 51 (33.4%) |  |
| Male | 225 | 141 (62.6%) | 84 (37.4%) | 0.061 |
| Female | 274 | 149 (54.3%) | 125 (45.7%) |  |

**Figure 2.**
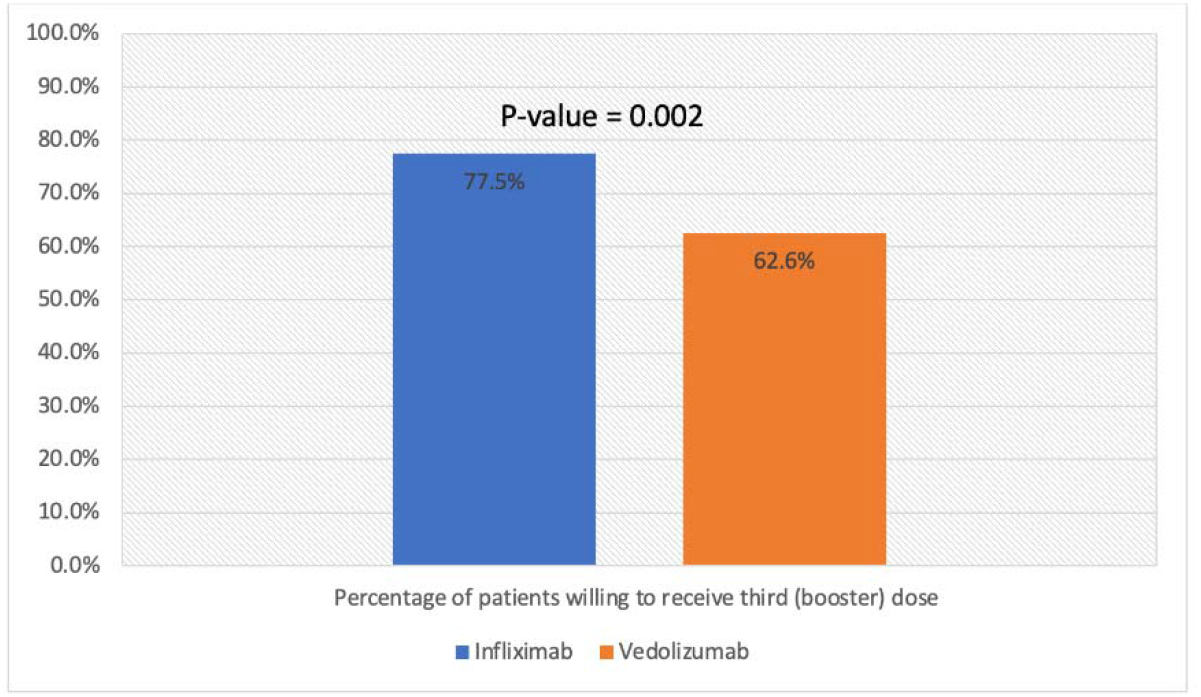
Percentage of patients willing to receive third (booster) dose stratified by biologic therapy.

## 4. Discussion

In this study, 499 patients with IBD were recruited to evaluate the willingness to receive third (booster) dose of BNT162b2 vaccine after receiving two doses of either BNT162b2 or ChAdOx1 nCoV-19 vaccines, which are the only available COVID-19 vaccines in Kuwait at the time of the study. Overall, 290 (58.1%) of the included patients received or were willing to receive the third (booster) dose. This is lower than the percentage of patients who received third (booster) dose among general population (77.5%).^15^ Similar to our study, a study conducted in Ontario, Canada, found that among patients with IBD, 58·3% had three doses as of Jan 9, 2022. Interesting, the percentage of patients who received third (booster) dose in patients with IBD was higher than the general population in Canada (44.3%).^16^

Another finding of this study was that patients who received two doses of BNT162b2 vaccines were more likely to receive a third (booster) dose compared to patients who received ChAdOx1 nCoV-19 initially. A possible explanation regarding the reduced willingness of patients to receive a third (booster) dose after vaccination with ChAdOx1 nCoV-19 vaccine could be explained by reports of thromboembolic events among people who had received the ChAdOx1 nCoV-19.^17^ Uptake of vaccines is highly dependent on public trust in the safety of vaccines, therefore, concerns of safety could be a possible reason of reluctance to get vaccinated.^18^ Another possible reason could be experiencing more adverse events after ChAdOx1 nCoV-19 vaccination. One study found that both vaccines induced transient side effects that ranged from mild to severe, however, ChAdOx1 nCoV-19 induced more severe side effects than BNT162b2.^19^ In addition, some patients who were vaccinated with ChAdOx1 nCoV-19 vaccine were reluctant to take a booster dose of BNT162b2 vaccine as they were concerned about the safety of mixing different type of vaccines.

In our study, there was no significant difference in the uptake of third (booster) dose among males and females. Unlike our study, gender was found to be one of the determinants of attitude towards COVID-1 vaccination in one study conducted in China.^20^ The study found that COVID-19 vaccination hesitancy existed in 50.7% of patients with IBD and merely 16.0% of participants opted for vaccination. The authors also found that the attitude of patients with IBD towards the third (booster) dose of COVID-19 vaccine was significantly more negative than that of the general population in China. Furthermore, 21.7% of patients with IBD considered the recommendations of their attending physicians as one of the three main reasons for getting vaccinated. Finally, the study demonstrated that safety and adverse events, as well as efficacy of COVID-19 vaccines are the top three concerns of patients with IBD for COVID-19 vaccination. We also found that patients with IBD treated with infliximab were more likely to get a third (booster) dose of COVID-19 vaccine compared to patients with IBD treated with vedolizumab. Possible explanation for this is that immunogenicity of patients treated with infliximab was attenuated after two doses of ChAdOx1 nCoV-19 or BNT162b2 vaccines.^21,22^ This may prompt patients to be more willing to receive a third (booster) dose. A study^23^ surveyed individuals with IBD to explore factors associated with vaccine uptake, concerns, and which sources of information were considered trustworthy surrounding vaccination. Authors found that self-perceived risk of being more unwell with COVID-19 due to IBD and past influenza vaccination, were positive predictors of COVID-19 vaccine uptake in IBD patients. Concerns about an IBD flare with vaccination is a unique consideration in those vaccine hesitant and is a negative predictor of vaccine uptake.

Another study^24^ assessed COVID-19 third (booster) dose willingness and hesitancy in Italian IBD patients, and investigated the determinants of these attitudes. Authors reported that more than 4 in 5 (80.3%) IBD patients were ready to receive third (booster) dose. Adherence to previous vaccinations had a positive influence on willingness to receive third (booster) dose. The study also observed positive association between vaccine willingness and a perceived higher risk of COVID-19 because of IBD. Finally, authors found that the most common reasons for COVID-19 vaccine hesitancy were the fear of adverse events and concerns about a perceived too-fast development.

Factors that increase likelihood of receiving third (booster) dose of COVID-19 vaccines in general population were explored extensively in the literature. One study^25^ measured factors associated with COVID-19 vaccine booster hesitancy and assess the roles of vaccine literacy and vaccine confidence in American adults’. A total of 2138 adult Americans aged 18 and over participated in this study. Among participants who were fully vaccinated or have received the first dose, an overwhelming majority (79.1%) would take a booster dose if recommended. In contrast, nearly half of the unvaccinated participants (46.3%) reported they would not take the booster dose. The study cited different factors that influenced this decision including socioeconomic, education level and demographic factors. In particular, those with a college education and graduate degrees were significantly more likely to accept third (booster) doses than those with only a high school education.

Furthermore, a questionnaire-based study was conducted in Algeria to determine COVID-19 vaccine booster acceptance and its associated factors in the general population.^26^ Authors found the most common reasons for acceptance were experts’ recommendations (24.6%) and the belief that COVID-19 vaccine boosters were necessary and efficient, while rejection was mainly due to the belief that primer doses are sufficient (15.5%), or that vaccination in general is inefficient (8%). Males, older individuals, those with chronic comorbidities or a history of COVID-19 infection, non-healthcare workers, and those with low educational levels were associated with significantly higher odds for booster acceptance.

Different international gastroenterological societies have published their recommendations regarding third (booster) dose of COVID-19 vaccination. The International Organization for the Study of Inflammatory Bowel Disease^8^ (IOIBD) recommends that all patients with IBD should receive any of the available COVID-19 vaccines as bad outcomes have been documented in patients with active IBD infected with SARS-CoV-2. The British Society of Gastroenterology^7^ (BSG) also recommends early vaccination for patients with IBD, as the benefits of vaccination outweigh the risks. Furthermore, the BSG recommends that immunomodulators should not be withheld for vaccination, and conversely, vaccination should not be deferred due to IBD medications. The Crohn’s & Colitis Foundation^27^ (CCF) largely agreed with the IOIBD. Additionally, the CCF was supportive of patients receiving additional doses if they were on immunosuppressive therapies. The CCF followed guidelines from the US Centers for Disease Control and Prevention stating that most patients with IBD, aside from patients on no medications or mesalamines only, qualified for an early booster.^28^ It is essential that Gastroenterologists are appropriately updated on efficacy and safety of the available COVID-19 vaccines as they play an important role in patient’s education as well as encouraging patients with IBD to get vaccinated. And currently, the effectiveness of the fourth dose of BNT162b2 is being investigated and new data is emerging regarding the best recommendations of the fourth dose.^29^

High level of B-cell immunity might be necessary to counterbalance the curtailment of cellular immunity effectiveness, which can be achieved by received a booster dose of COVID-19 vaccine. However, interestingly, there are some evidence that patients on anti-TNF therapy are less likely to get severe COVID-19. Because patients with IBD are already anxious about getting sick with COVID-19, it should be clear to them that although their cellular immunity maybe reduced with anti-TNF therapy yet they are not at increased risk of getting worse COVID-19 related outcome compared to the general population.^30^

The results of this study can be used to aid physicians, especially IBD specialists, researchers, and public health departments, to understand factors behind unwillingness to receive third (booster) doses develop, which in turn can be used to support creating measures and interventions to encourage patients with IBD to get a third (booster) doses. However, there are limitations to our study. Given that this is a cross-sectional observational study, selection bias and confounding variables are certainly one of the main challenges faced in observational setting. Therefore, as a result of the complexity of biases that can potentially affect an observational study, it is virtually impossible to conclude that our results are not due, in totality or in part, to different variables that cannot be controlled for. Examples of variables that could have affected the results are time and booster dose priority. It cannot be ruled out that with the passage of time, some patients may change their mind regarding willingness to receive the third (booster) dose. Also, at the initial phase of vaccination the third (booster) dose was available only to certain groups such as the elderly. Readers must carefully assess for possible explanations of an association. In addition, given the rapidly evolving nature of the pandemic, patients’ willingness for a booster dose may change over time. Finally, patients with IBD on other biologic therapies such as adalimumab and ustekinumab were not evaluated.

## 5. Conclusion

The percentage of patients with IBD willing or have received a third (booster) dose of COVID-19 vaccine was lower compared to general population. In addition, patients who received two doses of BNT162b2 vaccines were more likely to receive a third (booster) dose compared to patients who received ChAdOx1 nCoV-19. Patients treated with infliximab were more likely to receive a third (booster) dose of COVID-19 vaccine. Health education and recommendation from authoritative sources, such as government and IBD specialists, may support increase uptake of the third (booster) dose in patients with IBD.

## Supporting information

STROBE checklist

## Data Availability

All data produced in the present study are available upon reasonable request to the authors

## Supplementary Materials

STROBE guidelines (Table s1)

## Author Contributions

M.S.: Conceptualization, methodology, software, validation, formal analysis, investigation, resources, data curation, writing—original draft preparation, writing—review and editing. FA.: writing—original draft preparation, writing—review and editing. A.A.: validation, supervision, project administration. All authors have read and agreed to the published version of the manuscript.

## Institutional Review Board Statement

The study was approved by the standing committee for coordination of health and medical research of the Ministry of health of Kuwait (IRB 2021/1729) as per the updated guidelines of the Declaration of Helsinki (64th WMA General Assembly, Fortaleza, Brazil, October 2013) and of the US Federal Policy for the Protection of Human Subjects.

## Informed Consent Statement

Patients’ informed written consent was obtained before recruitment.

## Data Availability Statement

The data presented in this study are available on request from the corresponding author. The data are not publicly available due to legal and ethical restrictions.

